# Using the behaviour change wheel approach to optimise self-sampling packs for sexually transmitted infection and blood borne viruses

**DOI:** 10.1101/2021.07.06.21258646

**Authors:** P. Flowers, G. Vojt, M. Pothoulaki, F. Mapp, M. Woode Owusu, J. A. Cassell, C. Estcourt, J. Saunders

**Affiliations:** School of Psychological Sciences and Health, University of Strathclyde; Glasgow Caledonian University, Department of Psychology; Dept of Infection & Population Health, University College London; Department of Primary Care and Public Health, University of Brighton; Glasgow Caledonian University

## Abstract

**Purpose:** This paper describes the process of optimising a widely offered intervention - the self-sampling pack for sexually transmitted infections (STIs) and blood born viruses (BBVs). We drew upon the BCW approach, incorporating the theoretical domains framework (TDF) and the behaviour change technique taxonomy (BCTT) to systematically specify potential intervention components that may optimise the packs.

**Methods:** A behaviour change wheel analysis built upon prior thematic analyses of qualitative data collected through focus groups and interviews with members of the public and people recruited from sexual health clinics in Glasgow and London (n=56). Salient barriers and facilitators to specific sequential behavioural domains associated with wider behavioural system of pack-use were subjected to further analyses, coding them in relation to the TDF, the BCW’s intervention functions, and finally specifying potential optimisation in relation to behaviour change techniques (BCTs).

**Results:** Our TDF analysis suggested that across the overall behavioural system of pack use the most important theoretical domains were ‘beliefs about consequences’ and ‘memory, attention and decision-making’. BCW analysis on the overall pack suggested useful intervention functions should focus on ‘environmental restructuring’, ‘persuasion’, ‘enablement’, ‘education’ and ‘modelling’. Ways of optimising the intervention were also specified in relation to potentially useful behaviour change techniques (BCTs).

**Conclusions:** A detailed behavioural analysis building on earlier qualitative work using the TDF and the BCW provided a systematic approach to optimising an existing intervention. The approach enabled the specification of highly specific, evidence-based, and theoretically informed recommendations for intervention optimisation.

**What is already known on this subject?:**

- The use of self-sampling packs for sexually transmitted infections (STIs) and blood borne viruses (BBVs) has been widely implemented without in-depth assessment of user perspectives, adequate theorisation in relation to behaviour change, or optimisation
- In a previous qualitative study we reported on our use of thematic analyses (inductive and deductive) to understand the behavioural system of using self-sampling packs. We identified multiple modifiable barriers, and several important facilitators to using these packs, and their content, correctly
- Our thematic analyses showed that self-sampling packs offered a largely acceptable approach to STI and BBV testing and that with some modification it may be possible to increase both the range of people who can benefit from them and increase the return of samples

**What does this study add?:**

- This study theorises key barriers and facilitators to each sequential step within the behavioural system of using the self-sampling pack. Across the whole behavioural system of pack use we identified ‘beliefs about consequences’ and ‘memory, attention and decision-making’ as being particularly important theoretical domains
- To optimise self-sampling packs for STIs, our use of the behaviour change wheel suggested that to modify the pack we should use intervention functions that detail *environmental restructuring* and assist the user using pack contents through *persuasion* and *enablement* with some *education* and *modelling*
- The study provides an exemplar of how to use the behaviour change wheel within the process of intervention optimisation when building upon prior qualitative analyses. The results informed the redesign of the pack and the development of on-line support materials to be used within a large multi-site trial

## Introduction

Within this paper we show how, within the context of a large multi-site trial, we sought to systematically optimise an existing widely offered public health intervention (self-sampling packs for sexually transmitted infections (STIs)). Building on our previous qualitative analyses (Flowers et al, *3a*), here we report on our use of the behaviour change wheel approach (Michie, van Stralen, & West, 2011)(BCW) to systematically optimise self-sampling packs.

### The background to self-sampling packs

Across the UK home-based self-sampling packs for STIs and BBV are widely offered (e.g. Barnard et al, 2018; Ogale et al, 2019). Typically, a person requests a pack online, has it delivered to their home address, takes their samples (urine, genital swab, finger prick blood sample) at home, repackages up the kit and posts it back to the laboratory for testing. Results are provided by SMS (text message), telephone or email by the sexual health clinic.

The provision of self-sampling packs chimes with wider cultural changes clustering around the home delivery of a range of commodities, including diverse health technologies (e.g. COVID-19 self-sampling packs, bowl screening packs, or home-based genetic self-sampling kits). In light of recent changes to the ways people live and work wrought by COVID-19 it is likely that more and more health care will be delivered remotely. In the UK context, the widespread provision of self-sampling for STIs & BBV, sometimes to replace face-to-face options, has taken place at a time of austerity for many public services including sexual health. In some UK countries, it has also occurred at a time when the commissioning of sexual health services has been de-coupled from the National Health Service and repositioned within the remit of local authorities. In this paper, as part of a much larger sequential programme of work, we provide an example of how we used the BCW approach to optimise self-sampling packs for STIs in order to improve their use, enable their wider reach and increase sample return rate.

Our previous exploratory qualitative insights (Flowers et al, 2021a) highlighted problems with the existing self-sampling pack. Through thematic analyses with diverse members of the public and those attending sexual health services, we showed that that use of the pack overall was sometimes experienced as overwhelming, that pack-level instructions were challenging to many, and that the poor overall pack-usability was likely to reduce pack use.

As an already widely available and widely used intervention, the problems with existing packs do not prevent *all* people from using them effectively. Their problematic aspects may affect some people more than others. We know that pack users tend to be female, of white ethnicity and live in less deprived areas (Banerjee et al., 2018; Bracebridge, Bachmann and Ramkhelawon and Woolnough, 2012; Manavi and Hodson, 2017). Problems with the pack negate the pack’s role in reducing STI transmission because people cease engaging in sexual health self-care altogether, or drive them to attempt to use bricks and mortar sexual health services and through struggling to attain appointments are delayed or prevented from accessing timely treatment and wider care. It is also likely that the pack in its current form may be amplifying long standing inequalities and lead to the social stratification of sexual health screening and contribute to on-going sexual health inequalities (e.g. Wayal et al, 2017). Our earlier analysis suggested that those who are likely to struggle to use the pack are more likely to be people who have vulnerabilities such as low literacy, low health literacy, or low digital literacy (e.g. Middleton et al, 2021). Choices for sexual health care for these people may be reduced to only bricks and mortar clinics – and they may not be able to access them either.

Whilst these qualitative findings provided a clear, and user-led, sense of direction for considering ways of optimising existing packs, in themselves they did not offer a systematic way of considering how to improve the pack. Equally, the earlier qualitative work did not capitalise on the prior expertise and learning embodied within health psychological, or broader interdisciplinary, behaviour change theory or systematic approaches to consider ways of specifying intervention content to optimise the intervention. The BCW offers just such a systematic approach drawing on previous work. When the BCW is combined with the theoretical domains framework (Atkins et al., 2017; TDF) it offers a means to systematically borrow, and build upon, decades of previous research. As such, within this paper we focus on reporting how we systematically used these tools to develop highly specific ways of optimising self-sampling packs. Furthermore we draw upon the behaviour change technique taxonomy (BCTT) for similar reasons. The BCTT provides a common language to specify intervention content in terms of the indivisible techniques that can be harnessed to change behaviour. In this case the use of the self-sampling packs.

## Methods

### Participants

Details of the participants that generated the initial data that the current analyse can be found elsewhere (Flowers et al, 2021, paper 3a). Briefly, eleven focus groups and seven interviews with young heterosexual individuals and men who have sex with men (MSM) were conducted in Glasgow and London in late 2017. In total 56 participants took part in the study, of whom 25 were female and 31 were male. All were cisgender. Most participants were aged 18-25 years (n=36), identified as ‘white British’ or ‘white other’ (n=44) and were educated to University-level (n=39). Over half of the sample were heterosexual (n=40), with the remaining participants self-identifying as MSM, gay or bisexual (n=14). Around one third were recruited from sexual health services and had recently had a sexually transmitted infection, the others were sampled from the wider community. One quarter of the sample (n=9, 25%) had previously had an STI.

### Original Data collection

The primary data were collected within the focus groups and interviews using a topic guide and examples of the self-sampling pack (see Figure 1 below). As this study formed part of a programme of research into accelerated partner therapy, the process of identifying, testing and treating sex partners of people with STIs, lustrum.org.uk, the pack also contained antibiotics. The topic guide was used to steer discussion across the full range of issues detailed; where possible within the focus groups, discussion between participants was encouraged rather than between facilitators and participants. The pack contained self-sampling kits for vulvo-vaginal swabbing, urine collection and taking finger prick blood samples. The pack included a variety of information sheets giving instructions on how to use the kits, a return envelope and a series of labels to adhere to completed samples.

**Figure 1:**
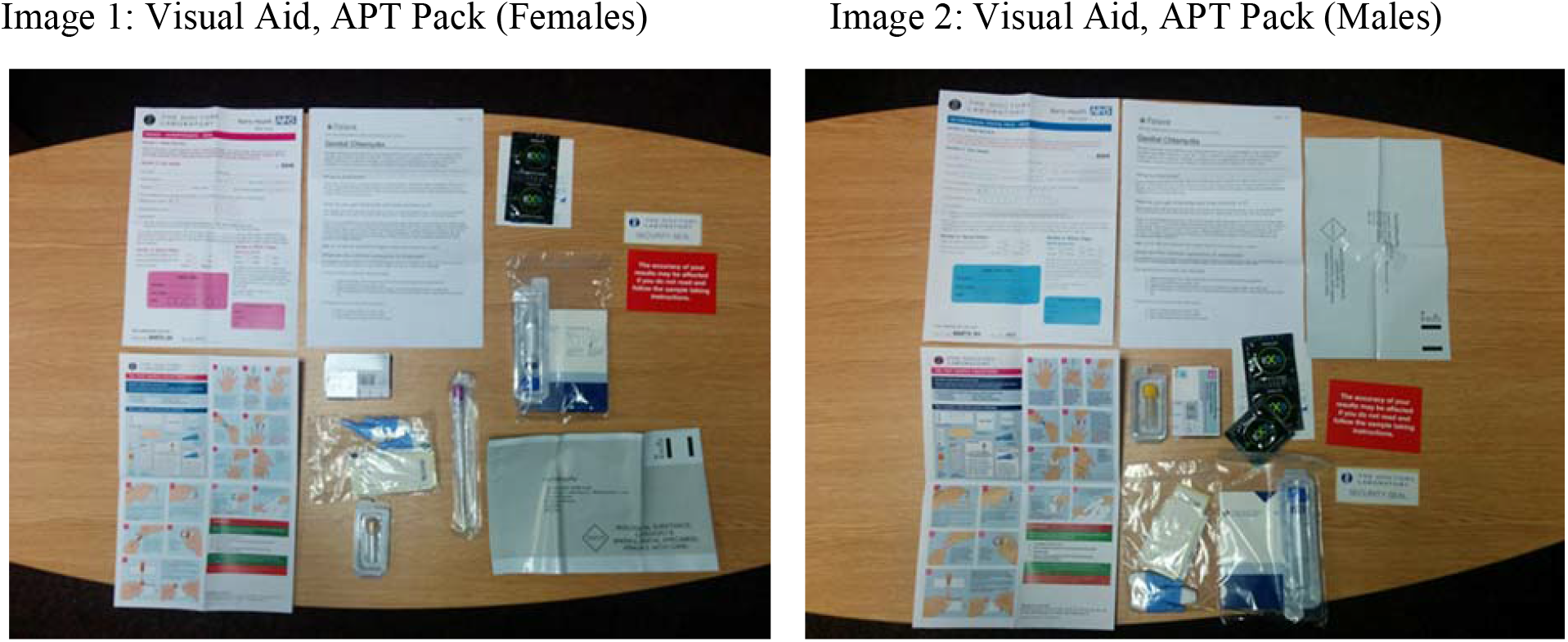
Images of the pack and its contents used within initial data collection.

### Data analysis

The original thematic analysis is reported elsewhere (please see Flowers et al, 2021, paper 3a for details)

In developing, or optimising, behaviour change interventions two key principles are important; the notion of parsimony and the idea of behavioural specificity. Parsimony is about offering the simplest and most elegant explanations of often complex phenomena. Behavioural specificity, on the other hand, highlights that that the best insights into behavioural change occur when we can harmonise the highly specific nature of our understandings of antecedents to behaviours with an equally fine grained, highly specific, understanding of the behaviour that we seek to change (e.g., see Presseau et al., 2019).

For us, as a pragmatic and applied interdisciplinary team, we have sought to attain the best balance of both behavioural specificity and parsimony. With regard to considering the behaviours at the heart of our behaviour change endeavour, having conducted inductive, participant-led thematic analysis detailing how our participants thought about the pack and what constituted pack use, we conceptualised using the self-sampling packs as a behavioural system structured by a series of inter-related and sequential behavioural domains (see Flowers et al., 2021, paper 3a). Although we could have analysed each behavioural domain in even more specific behavioural terms, we felt it was very likely that the number of shared antecedents for these highly-related specific behaviours would be high and that there could be a lot of effort for minimal reward. Equally we could have tried to identify a single behaviour within the system-chosen because of its potential for wide ranging spill over effects. Again, however, because of the sequential nature of many of the behaviours and because the pack was already widely used this did not seem to be the optimal approach.

Figure 2 illustrates how we conceptualised pack use; conceptualising sequential elements as distinct behavioural domains within an overarching behavioural system. It is worth noting that effective pack use requires completion of all four behavioural domains within this behavioural system. However, the specificity of intervention optimisation demands distinct behavioural analyses at the level of each domain.

**Figure 2:**
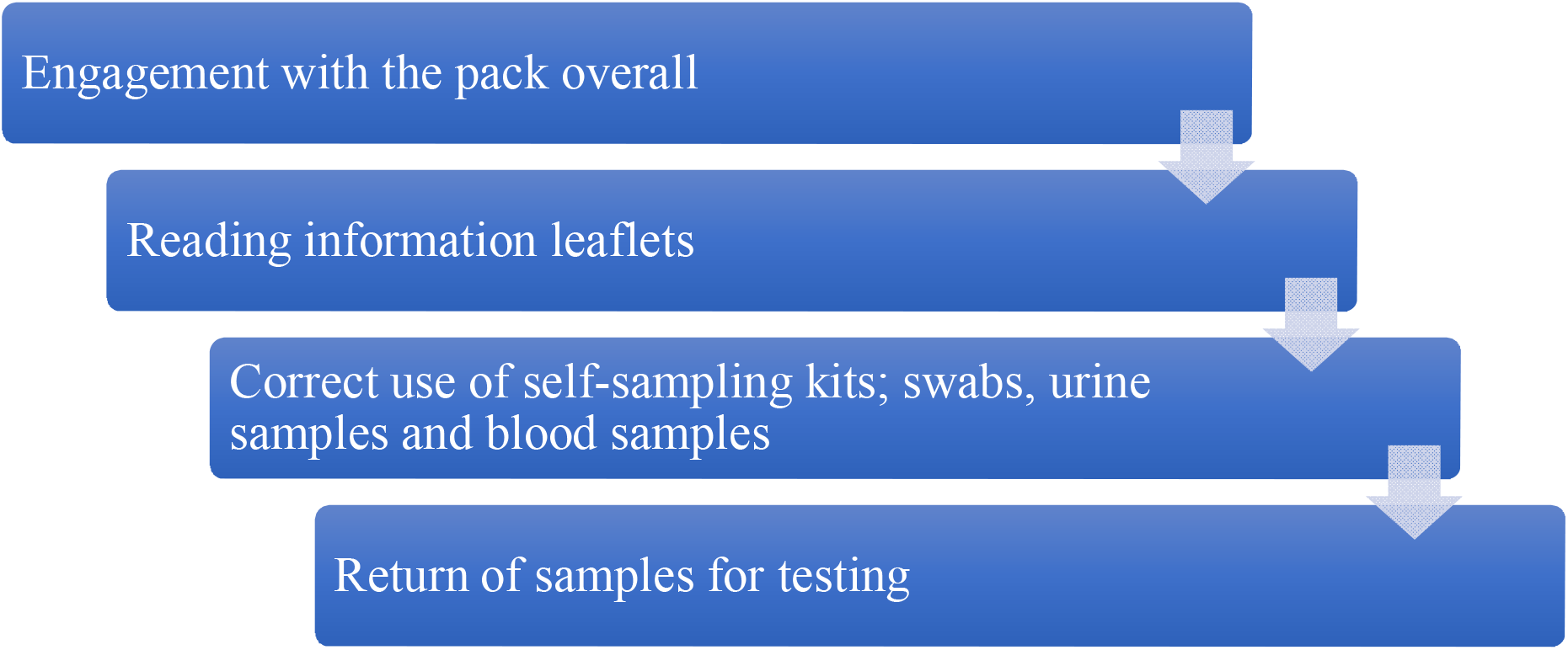
Balancing parsimony and behavioural specificity to illustrate the behavioural system of using self-sampling packs and associated behavioural domains

For the applied and pragmatic focus of the work reported here, and for the goals of this project, Figure 2 captures a useful degree of behavioural specificity to enable intervention optimisation. In relation to finding a pragmatic, parsimonious approach to understanding the determinants of our behavioural domains, one the one hand, the plethora of explanatory frameworks that theorise behaviour change within traditional health psychology models and theories are too fine grained. Yet, on the other hand, popular approaches such as the Capability, Opportunity and Motivation model of behaviour change (COM-B model; Michie, Atkins & West, 2011) categorises the antecedents of behaviour at too broad a level, only providing explanations of behaviour change within six broad categories. As such, for this project, we felt TDF offered a suitable, mid-level, explanatory framework that enables a good level of specificity in relation to understanding the casual mechanisms underpinning behaviour change within the sequential behavioural domains that constitute the overall behavioural system of pack use.

In summary we believe we have found a good balance of parsimony and specificity in relation to the process of behavioural diagnosis. This plays out at two levels; firstly, identifying the right *pitch* of our behavioural foci for intervention optimisation; and secondly, in relation to theorising the causal mechanisms that shape these behaviours at a useful level (i.e. the TDF rather than COM-B).

### The theoretical domains framework

The TDF is a meta-theoretical framework that integrates 14 key theoretical domains known to be important in understanding behaviour change across a range of populations and settings and health arenas (Cane et al., 2012; Atkins et al., 2017). Its domains provide a relatively high-level common language to describe clusters of theoretical constructs that shape behaviour. As such we refer to them as causal mechanisms (i.e., they describe the *causes* of behaviour within the language of multiple theories). In other words, the TDF’s domains provide a coherent and structured way of organising explanations of why behaviours, such as those involved in the effective use of a self-sampling packs, either *do*, or *do not* occur.

The TDF’s domains are domains because they de-duplicate, synthesise, and summarise a myriad of multiple previous theoretical constructs from formal theoretical models of health psychology. In this way, the TDF provides a high-level theoretical framework to describe the range of causal mechanisms implicated in understanding behaviour and behaviour change. Critically, the TDF can be used as a precursor to subsequent wider intervention development processes, using for example, the complementary meta-theoretical framework - The Behaviour Change Wheel (BCW) (Michie, Atkins & West, 2011). Here we use it in the process of intervention optimisation. Figure 3 illustrates how the 14 domains of the TDF map and structure the causal mechanism shaping behaviour (the specific behaviour being changed is at the centre of the circle).

**Figure 3.**
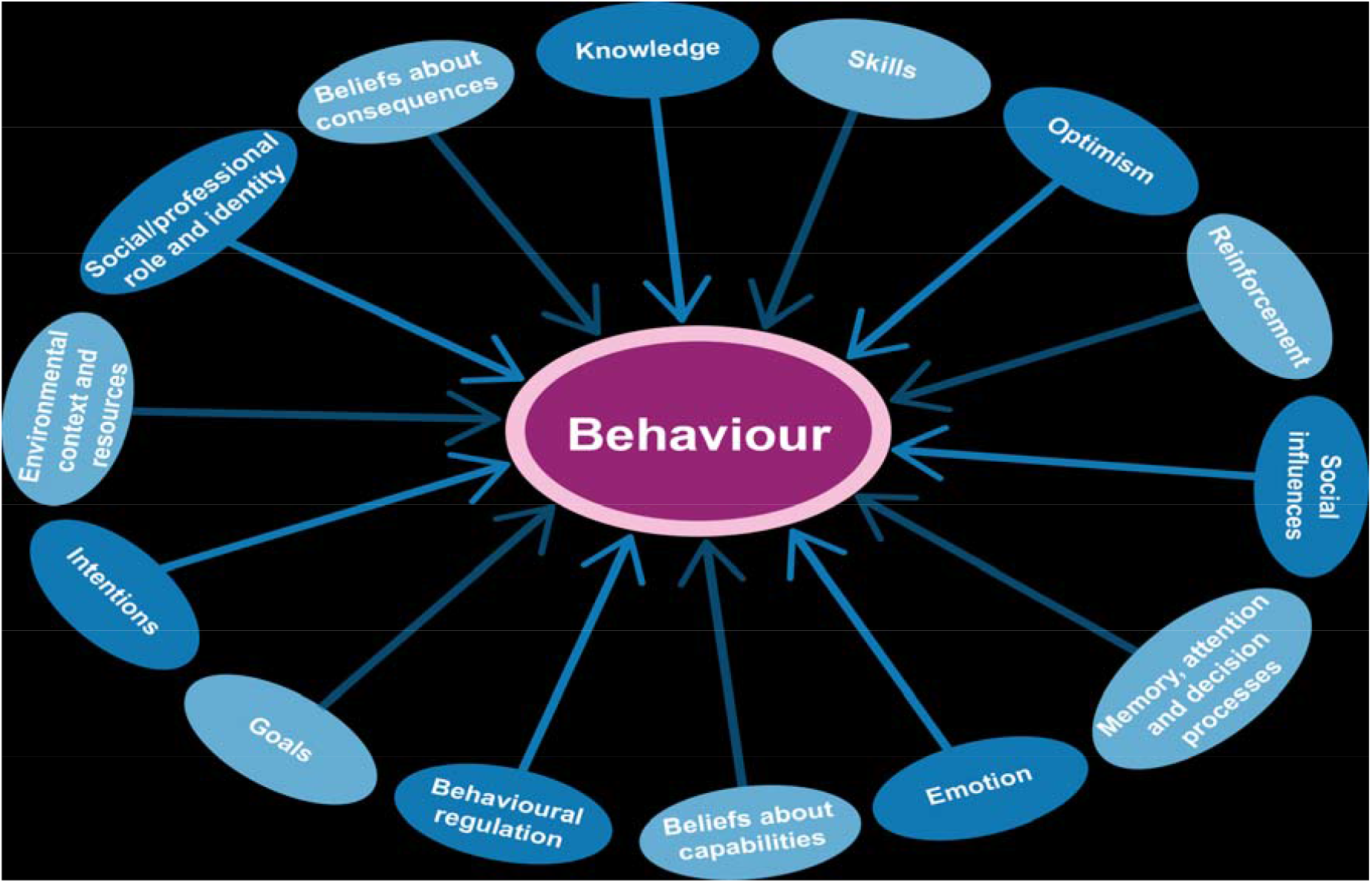
The theoretical domains framework mapping the casual mechanisms that can moderate any specific behaviour.

### The behaviour change wheel approach

The BCW like the TDF, is also an over-arching meta-framework representing the synthesis of many other approaches to behaviour change. The BCW synthesises 19 behaviour change frameworks all designed for the development and evaluation of behaviour change interventions (Michie et al., 2011). The BCW details nine intervention functions that represent broad categories by which interventions can change behaviour: ‘Education’; ‘Persuasion’; ‘Incentivisation’; ‘Coercion’; ‘Training’; ‘Enablement’; ‘Modelling’; ‘Environmental Restructuring’; and ‘Restrictions’. These are prescriptively mapped to the specific components of TDF, or alternatively, the COM-B. In this way, future intervention functions are tailored to demonstrable intervention need (e.g., the causal mechanisms specifically identified through prior TDF analysis).

### The behaviour change techniques taxonomy

As part of the wider behaviour change wheel approach, Michie et al (2013) developed an extensive 93-item list of behavioural techniques, which was consensually agreed regarding impact and importance, and where every technique was clearly defined and exemplified. A behaviour change technique is a specific irreducible clearly defined component of an intervention. It is directly related to larger ‘intervention functions’ and therefore with the causal processes underlying behaviour change (i.e. TDF). For example, if the intervention function was ‘modelling’, then an associated behaviour change technique that could be part of the operationalisation of this function could be ‘demonstrating the behaviour’.

### How these tools were used within this paper

Our use of the behaviour change wheel approach to intervention optimisation centred on a three step approach. These three steps were initially conducted by PF, an HCPC registered Health Psychologist and then audited by two other psychologists (MP and GV), all of whom are expert and actively using the BCW approach, before discussing with the wider interdisciplinary team (medical clinicians, public health professionals, health service researchers, health economists). Throughout we aimed to do justice to these conceptual tools building on their synthesis of prior expertise and learning but also to draw upon our subject-specific expertise, the context of the work and the data we had collected.

**Step 1:** For the current paper, our previous work identifying the barriers and facilitators to the range of behavioural foci within the overall system of using the self-sampling pack was further analysed initially using the TDF (Cane et al., 2012). Each identified barrier or facilitator could be coded against multiple TDF domains. The relative frequency of particular TDF coding was assumed to be indicative of the relative importance of the causal mechanism it presents at the level of each behavioural domain but also in relation to considering the wider behavioural system.

**Step 2:** The TDF domains were further analysed using the BCW approach (Michie et al., 2014). Here, TDF domains were matched prescriptively with appropriate intervention functions for each barrier and/or enabler (see Table A in supplementary files).

**Step 3:** The relevant intervention functions were further examined, and working iteratively with the previous qualitative analysis to tailor BCTs to this particular context, we specified and operationalised potentially useful behaviour change techniques (BCTs) to specify useful optimised intervention content.

### Research questions

1. How can the barriers and facilitators to the range of behavioural domains within the behavioural system of pack use for STI and BBV self-sampling *be theorised* using the TDF?
2. What potentially useful *intervention functions* and potentially useful *behaviour change techniques* can be specified and operationalised using the BCW to optimise self-sampling packs for STI and BBV testing?

## Results

**Table A** in the supplementary file shows the results of the complete BCW analysis. It can be read both vertically and horizontally. Reading vertically from top to bottom illustrates the various sequential behavioural domains associated with the behavioural system of effective use of the pack (as shown in *Figure 1*). Reading horizontally, left to right, across the table it is possible to see the target behaviour (pitched at the level of the behavioural domains) at the heart of the BCW analysis that is to be changed (left hand column). Secondly, we present the specific barriers and facilitators to changing that behaviour summarising insights from our earlier qualitative work (Flowers et al., 2021, paper 3a). Thirdly, we show how these barriers and facilitators (as causal influences on the enactment of each behavioural domain) were coded in relation to theoretical domains specified by the TDF. Fourthly, we show how the TDF domains, when matched with the elements from the BCW, specify broad intervention functions that correspond to the underlying barrier or facilitator. Finally, at the far right we show how these intervention functions can be expanded and detailed in relation to the specification of particular behaviour change techniques that should optimise the intervention. The optimisation process works because i) it is based on the evidence we collected through our earlier qualitative work; and ii) it draws on the decades of earlier theoretical work encompassed within the TDF; iii) it uses the standardising language of the intervention functions from the BCW approach and iv) it uses the specific language of the BCTT. The interested reader should be able to gauge the logic of each row independently and see how we provide relatively granular solutions to the behaviour change problems identified by our participants within our previous qualitative work.

### 1) How can the barriers and facilitators to the range of sequential behavioural domains associated with the behavioural system of pack use for STI and BBV self-sampling be theorised using the theoretical domains framework?

In this section the results of our TDF analysis are presented within a narrative format that should be read alongside Table A within the supplementary file. Table 1 shows the relative frequency of particular TDF domains for each of the four sequential behavioural steps.

**Table 1.**
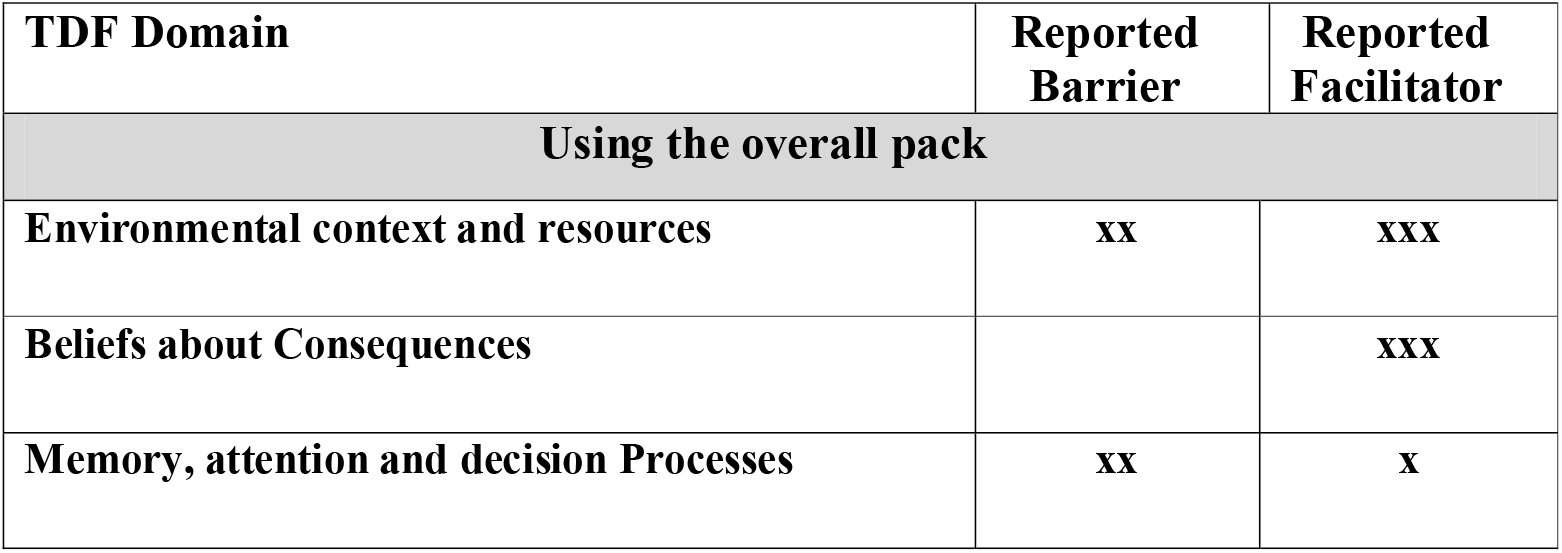
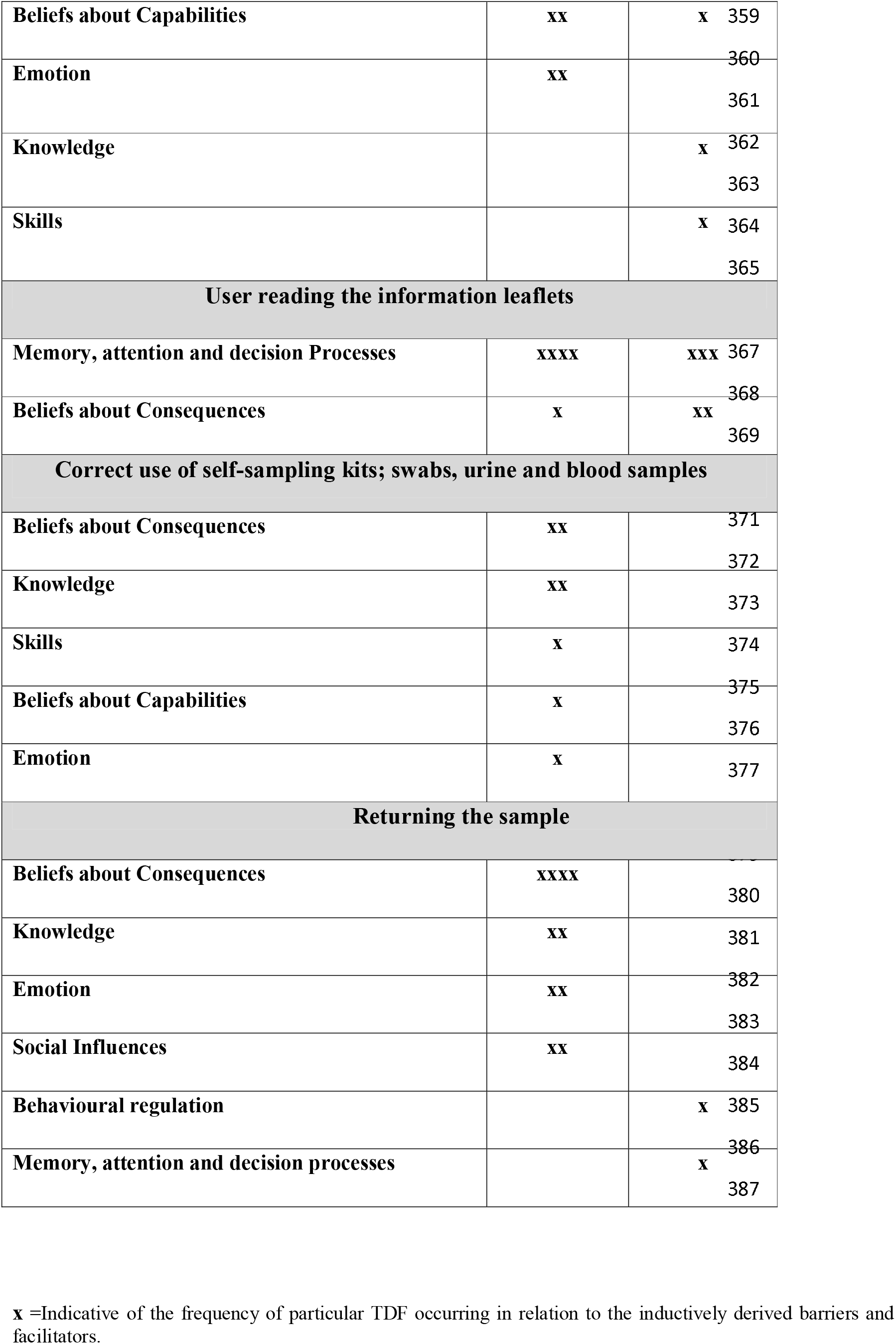
Causal mechanisms relevant for each behavioural domain within pack use.

Table 1 shows that for the broad behavioural domain of ‘*the user using the overall pack’* the pack overall created a particular environmental stressor presenting significant anxiety and wider challenges to users’ beliefs about their capabilities in using the pack. These were understood to challenge users’ attention and lead to potential cognitive overload. In contrast to these reported barriers, pack use was understood to be facilitated by the user’s appreciation of the consequences (i.e. relative ease) of using the pack (despite its complications) when compared to visiting a clinic and the negative sequelae of doing so. Procedural knowledge concerning how to use the overall pack and a revised compartmentalisation of the pack content were also suggested as ways overall pack use could be enhanced.

For the step of ‘*reading the information leaflets’* the key casual mechanisms implicated were the user’s reduced attention and associated ability to follow the specific instructions related to the formatting of leaflets. However, our participants also suggested the inclusion of information about the consequences of untreated infections at the start of the leaflets might increase attention whilst reading the leaflets. In turn this might enable the user to read the somewhat challenging level of information contained within the leaflets. Equally, modifying the leaflets to be clearer was thought to enhance attention and decision-making. Moreover, the inclusion of information about how long it might take to complete the task was also suggested as a means to enhancing reading the full leaflets.

In relation to important causal mechanisms implicated in the *correct use of self-sampling kits*, these included knowledge and skills deficits and low perceived ability to use the kits themselves. Knowledge deficits included both erroneous beliefs about the potential for samples to degrade in the post, an outdated and spurious understanding of HIV and the implications of a positive diagnosis. Beliefs about capabilities related to poor perceived self-efficacy in relation to collecting samples. No corresponding facilitators were recorded.

In relation to the behavioural domain of the pack, user *posting their sample back to the labs for processing* Table 2 shows an overview of the key causal domains that are important in explaining this behaviour. There were key concerns about the return envelope being recognised by others as being related to sexual health screening or a diagnosis of an STI and in this way STI-related stigma may prohibit return of the samples. Furthermore, there were concerns that the postal system may be inadequate (on a number of levels) in ensuring the safe return of samples and in turn this may decrease the likelihood of people returning the samples within the prepaid envelope. Participants believed that return of the samples could be enhanced with the provision of multiple ways of returning packs (e.g. drop off in a pharmacy), ensuring decision-making processes could tailor user choice of the way to return the samples that felt safe for them. Moreover, it was also suggested that processes which notified the user of safe receipt of their sample may also be useful.

**Table 2.**
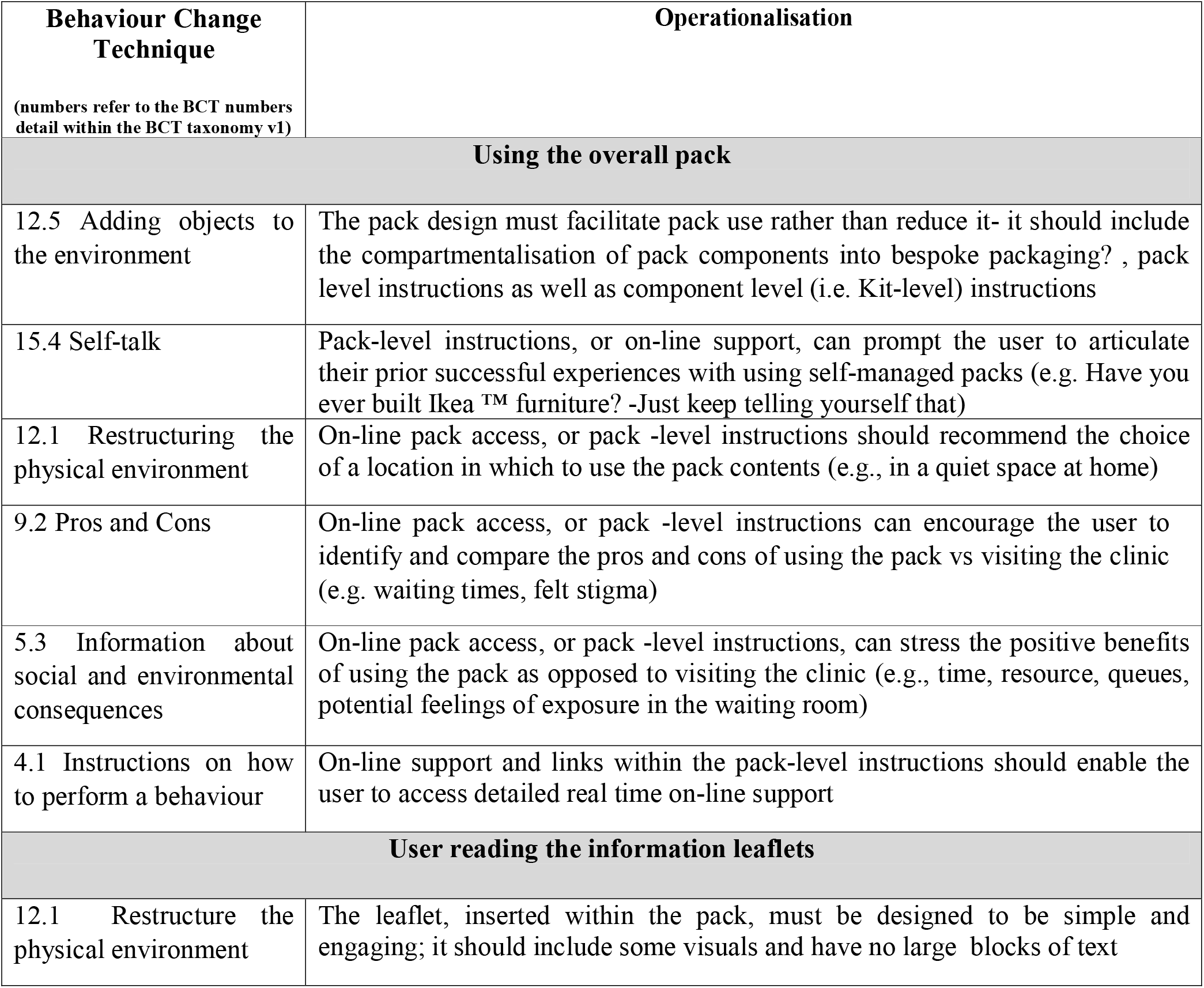
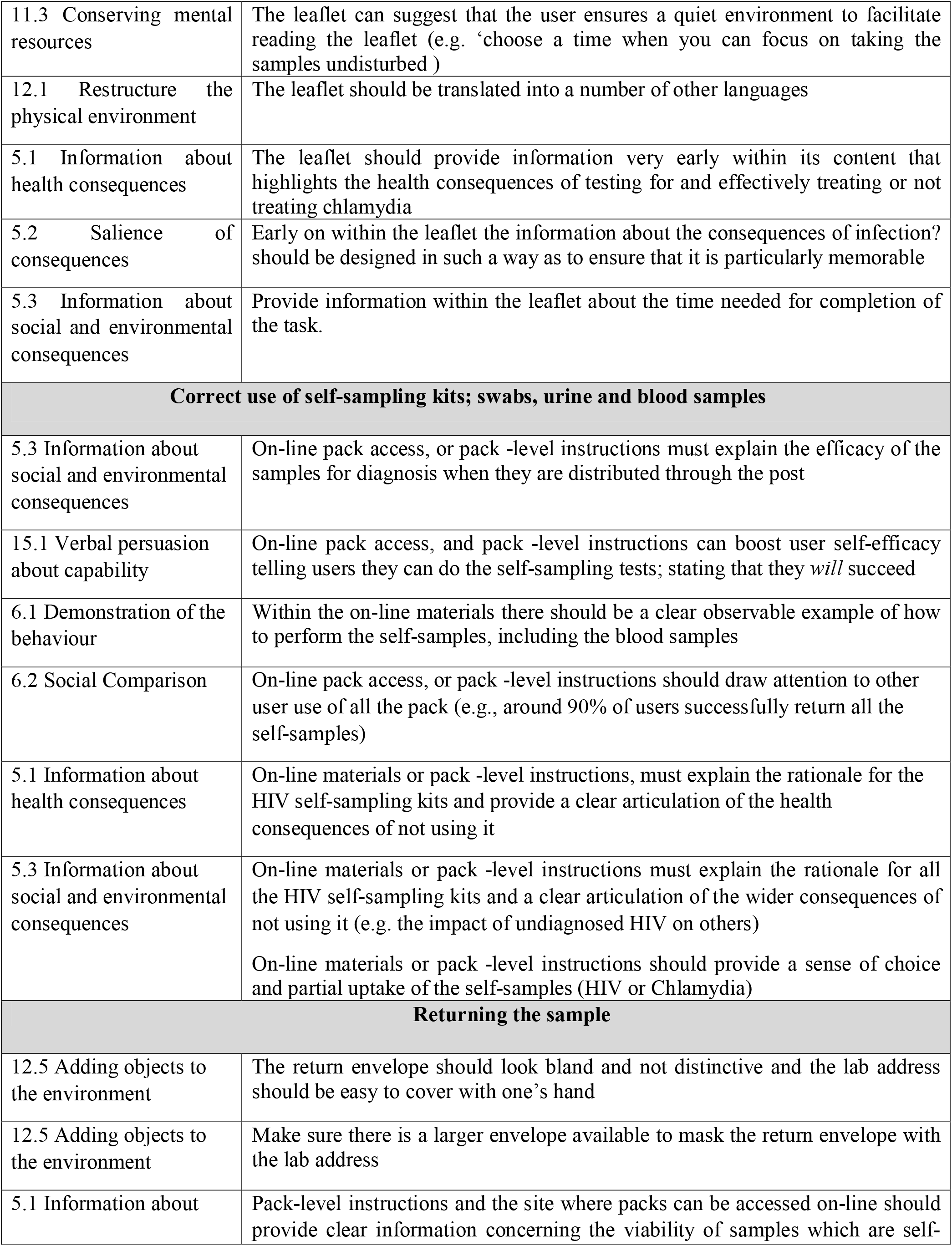
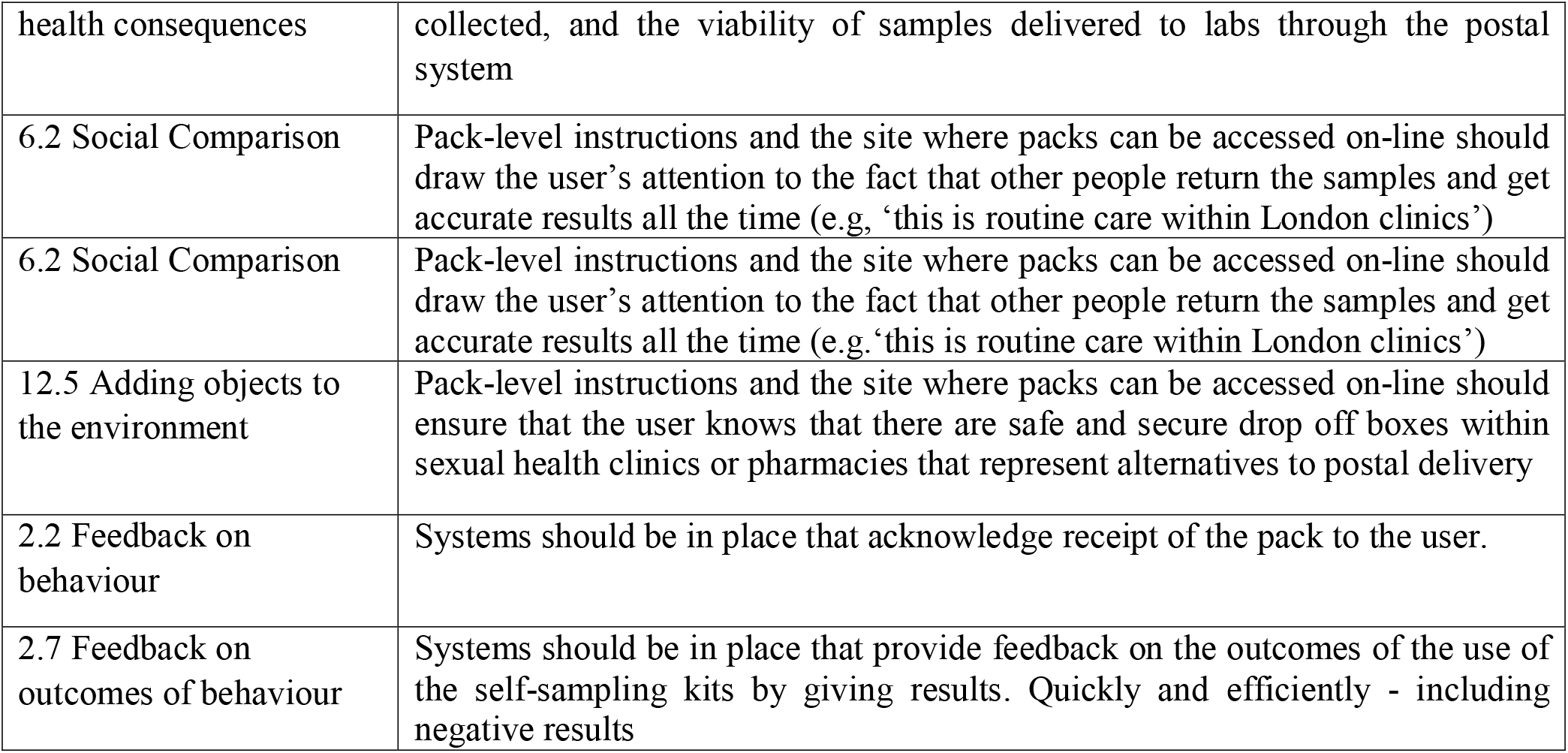
Suggested intervention functions and behaviour change techniques to optimising behaviour hange in relation to each behavioural domain within pack use.

### 2. What potentially useful intervention functions and potentially useful behaviour change techniques can be specified using the behaviour change wheel (BCW) to optimise self-sampling packs for STIs and BBVs?

Again, we draw the reader’s attention to Table A within the supplementary file which shows the full matrix of our initial findings concerning the barriers and facilitators to completing each behavioural domain within the overall behavioural system of pack use, the relevant TDF domains identified in the step above, and the corresponding intervention functions in addition to examples of how they could be operationalised as individual behaviour change techniques (BCTs). In this way, each row within Table A provides an auditable account of our use of these tools for the whole process of intervention optimisation. Within each row there is a sense of how we have ‘reverse engineered’ potential intervention content in ways that are precisely tailored to the findings from our earlier qualitative work and the causal mechanisms identified through the use of the TDF. Within this section however, to address the research questions directly, we present a narrative account of our findings concerning intervention functions and then tabularise our suggestions for potentially useful BCTs.

To address the behavioural domain of ‘using the overall pack’ our analysis suggested intervention functions should focus on *environmental restructuring* through modifying the pack and providing on-line materials to *enable* pack use. On-line materials should also *model* ideal pack use (for example, depicting pack use in a quiet location within online materials) and *persuade* the potential user that using the pack is a better way of testing than trying to get an appointment within a clinic and that they can overcome perceived barriers to pack use.

For the behavioural domain of ‘reading the information leaflets’ our analysis suggested that useful intervention functions should focus on *environmental restructuring* through changing the leaflets to improve accessibility and making sure the user is reading the leaflets within a quiet place. The leaflets should also be modified to *persuade* and *educate* the reader around the importance of reading the leaflet and following its instructions.

For the behavioural domain of ‘Correct use of self-sampling kits; swabs, urine and blood samples’ intervention functions are required that focus on *environmental restructuring* that provide additional on-line support that *educates, models, trains* and *persuades* the viewer to use the sampling kits correctly. Barriers to Kit-users can be overcome through persuasion concerning the consequences of kit use and benchmarking others normative behaviour around kit use.

Finally, for the behavioural domain of the user returning the samples useful intervention functions could use *environmental restructuring* through modifying the pack and specifically the return envelope to *enable* users to overcome perceived barriers. Furthermore, users can be *educated* and *persuaded* about the efficacy of the postal system and the labs in processing accurate results from samples through the modification of pack level content. Finally, intervention functions that *educate* and *enable* those returning samples about the receipt and results of their sampling may be useful modifications.

Overall, across the whole behavioural system, our analysis of potentially useful intervention functions shows that environmental restructuring (n=16), persuasion (n=14), enablement (n=14), modelling (n=9), education (n=9) and training (n=1) could all be important in optimising the pack.

Table 2 illustrates the end product of our analyses and details the way behaviour change techniques could be operationalised which reflect intervention functions and specify evidence-based and theoretically informed ways of optimising the intervention. We would recommend all the suggested changes to optimise the self-sampling packs.

Our analysis shows that different sequential behavioural domains that make up the overall behavioural system of pack use warrant distinct behaviour change techniques. However, in relation to describing the overall frequencies of recommended BCTs across the whole behavioural system, these related to the BCT grouping ‘natural consequences’ (n=10), ‘antecedents’ (n=9), ‘comparison of behaviour’ (n=4), ‘feedback and monitoring’ (n=2), ‘comparison of outcomes’ (n=2), ‘self-belief’ (n=2), ‘shaping knowledge’ (n=1), and ‘regulation’ (n=1).

## Discussion

This paper presents the results of a behaviour change wheel analysis of a prior thematic analysis of qualitative data collected through focus groups and interviews. It presents the granular specification for changes that should optimise an existing intervention using concepts and language from the BCW; theoretical domains specified by the TDF, intervention functions, and the behaviour change technique taxonomy. Our analysis focused upon ways to optimise an existing individual-level intervention. The existing intervention itself can be conceptualised as relatively simple when compared to interactional or longer term continuous interventions. It includes the effective use of a pack that contained several self-sampling kits, instructions and materials to enable the return of samples to lead to diagnosis.

Recent studies however, show that return rate of samples is highly variable and socially patterned. The use of self-sampling packs is likely to be of increasing importance remote and self-managed care becomes more and more normative. Self-testing / sampling for COVID-19 is also a central tool to returning to the work or educational setting. Insights into how to optimise the use of such packs are likely to be increasingly useful. To our knowledge this paper is the first internationally to focus on the optimisation of such self-managed packs using tools from health psychology and the wider behavioural sciences.

Our use of the behaviour change wheel may be interesting to other health psychologists in the way we sought to balance behavioural specificity with parsimony. The effectiveness of the BCW approach relies on rigorous examination and selection of the appropriate specific behaviour(s). In our experience, this first step in the process of a ‘behavioural diagnosis’, in other words selecting the key behaviour(s) to be changed within given behavioural domains and their wider systems, is far more complex than published guidance and papers would suggest, yet it is central and indeed fundamental to the usefulness of the whole BCW approach. Michie et al (2014) for example highlight the importance of considering likely impact, ease of implementation, likely spill-over and ease of measurement. However, for an intervention already existing and known to be less than optimal this approach with its singular focus was deemed insufficient to enable the overall optimisation that would lead to wider population reach and effective use. As such we cast our behavioural lens across the whole behavioural system of pack-use.

Equally of interest to health psychologists and other behavioural scientists we spent a lot of time considering how to conceptualise the diverse behaviours involved within effective pack use. We believe it is important to stress to others the centrality of this step in relation to using tools like the BCW approach effectively. Within this study we sought to conceptualise the overall use of the pack as a behavioural system; for us a behavioural system is a series of interdependent behavioural domains, each domain in turn is constituted from a number of discrete behaviours. We made the decision to pitch our analysis at the level of the behavioural domain and not the overall behavioural system or indeed at the level of highly specific behaviours. Our findings show that there are important differences in the causal mechanisms (i.e., TDF domains) associated with the each of the different behavioural domains and this resulted in us specify different intervention content for each sequential step. We believe that there is a need for more conceptual and methodological work in this area as many papers using these tools fail to provide clarity about this essential first step in behavioural diagnosis.

Accordingly, having established a pragmatic way of conceptualising the behaviours we wished to optimise, using the BCW, we were able to make distinct and quite granular recommendations for ways to intervene in each behavioural domain. By quantifying the overall frequency of the TDF domains we coded as indicative of their relative importance, we were able to evidence that optimisation should focus overall on changes to the user’s ‘*beliefs about consequences*’ when using the pack, and additional information to facilitate the user’s ‘*memory attention and decision processes’*. We also provided particular detail of ways to optimise each of the behavioural domains involved in this wider behavioural system. Subsequently, we drew upon the wider team’s clinical, practical and inter-disciplinary expertise and conducted an APEASE analysis of the recommendations developed here. APEASE addresses affordability, practicability, effectiveness/cost-effectiveness, acceptability, side-effects/safety and equity.

Our final analysis and the agreed BCTs and intervention content were eventually used to develop a series of on-line support videos for people using the packs. Our analysis was also used to redesign the packs that were used within a multi-site cluster randomised control trial of APT.

## Strengths and limitations

*Limitations* of the current study included the potential for recruitment bias. Although we recruited participants from both clinical and community settings and in Scotland and England the participants that agreed to take part were largely well educated, health literate and confident and/or comfortable enough to share their views and opinions on self-sampling packs for sexually transmitted infections. In this way our insights into optimising self-sampling packs are based on the perspectives of participants who may not represent the people for whom optimisation may well be most important. In light of this we also conducted similar research amongst people with mild learning difficulties to try to provide insights from more diverse populations (Middleton et al., 2021). Overall we found similar barriers and facilitators to those reported in Flowers et al., 3a).

*Strengths* of the current study are that the optimisations are all based on empirical qualitative work and have benefited from the use of behaviour change theory. We have presented a clear account and auditable summary of our decision-making and the interpretation of results (Table A in appendix). Other strengths include the fact that our results have been translated into a range of resources that have been available to optimise self-sampling packs for STIs. In addition, we have provided a transparent account of using behavioural theory to optimise an existing intervention.

## Conclusion

Using tools from behavioural science and health psychology we have shown how it is possible to systematically optimise STI & BBV self-sampling packs to increase their reach and effective use. Whilst our analysis here focused upon optimising self-sampling packs for use within a wider trial of a particular form of partner notification we believe our analysis offers several transferable insights into self-sampling packs more generally and for self-sampling packs for COVID -19 in particular.

## Data Availability

Data available upon reasonable request to the corresponding author.

## Notes

### Competing Interest Statement

The authors have declared no competing interest.

### Clinical Trial

RP-PG-0614-20009

### Clinical Protocols

https://bmjopen.bmj.com/content/10/3/e034806

### Funding Statement

This work presents independent research funded by the National Institute for Health Research (NIHR) under its Programme Grants for Applied Research Programme (reference number RP-PG-0614-20009).

### Author Declarations

Ethical approval from Glasgow Caledonian University Research Ethics Committee (HLS/PSWAHS/A15/256) and NHS Ethics Approval (16/NI/0211) were obtained.

