## Supplemental Table A for "Using the behaviour change wheel approach to optimise self-sampling packs for sexually transmitted infection and blood borne viruses"

| **Supplementary file: Table A**  **‘Using the behaviour change wheel, incorporating the TDF and BCTT, to specify recommended ways of optimising self-sampling packs’** | | | | | |
| --- | --- | --- | --- | --- | --- |
| **Behavioural domain to be moderated** | **Barriers** | **Facilitators** | **Mechanism of Action (TDF domain)** | **Intervention Function** | **Recommended intervention content including Behaviour change techniques (indicated by number) to be used within interventions** |
| **Pack user uses the overall pack** | For the pack user, the pack can look overwhelming and chaotic and can put off overall engagement |  | Environment, context and resource  Emotion  Memory, Attention and decision processes  Beliefs about capabilities | Environmental restructuring  Persuasion, modelling, enablement | 12.5 Adding objects to the environment  The pack design must facilitate pack use rather than reduce it- it should include the compartmentalisation of pack components, pack level instructions as well as component level (i.e. Kit-level) instructions |
|  | Pack-related stress can reduce attention and engagement with the pack and its contents |  | Emotion  Memory, Attention and decision processes | Persuasion, modelling, enablement | 15.4 Self-talk  Pack-level instructions, or on-line support, can prompt the pack user to articulate their prior successful experiences with using self-managed packs (e.g. Have you ever Built Ikea furniture? -Just keep telling yourself that you can do it) |
|  | The individual circumstances of pack users might add to problems with pack use (e.g., youth or sharing accommodation) |  | Environment, context and resources  Beliefs about capabilities | Environmental restructuring  Modelling  Enablement | 12.1 Restructuring the physical environment  On-line pack access, or pack -level instructions should recommend the choice of a location in which to use the pack contents |
|  |  | Pack user understanding of the relative ease of using the pack compared to the visiting the clinic could enhance overall engagement with the pack | Beliefs about consequences | Education, Persuasion  Enablement | 9.2 Pros and Cons  On-line pack access, or pack -level instructions can encourage the pack user to identify and compare the pros and cons of using the pack vs visiting the clinic (e.g. waiting times, felt stigma)  5.3 Information about social and environmental consequences  On-line pack access, or pack -level instructions can stress the positive benefits of using the pack as opposed to visiting the clinic (e.g., time, resource, queues, potential feelings of exposure in the waiting room) |
|  |  | For the pack user a weighing- up of the potential stigma and embarrassment associated with visiting the clinic compared to using the pack can facilitate pack use | Beliefs about consequences  Memory, attention and decision-making | Education, persuasion, modelling | 9.2 Pros and Cons  On-line pack access or pack level instructions can encourage the pack user to identify and compare the pros and cons of using the pack vs visiting the clinic  5.3 Information about social and environmental consequences  On-line pack access or pack level instructions can stress positive benefits of using the pack as opposed to visiting the clinic (time, resource, queues, potential feelings of exposure in the waiting room) |
|  |  | Inclusion of pack-level instructions could support engagement with the pack | Knowledge  Beliefs about capabilities  Beliefs about consequences | Persuasion  Modelling  Enablement | 12.5 Adding objects to the environment  The pack design must facilitate pack use rather than reduce it- this should include compartmentalisation of pack components, pack level instructions as well as component level instructions |
|  |  | A clear physical framework that organised the pack contents could support engagement with the pack | Environmental context and resources | Environmental restructuring  Persuasion  Enablement | 12.5 Adding objects to the environment  The pack design must facilitate pack use rather than reduce it- this should include compartmentalisation of pack components, pack level instructions as well as component level instructions |
|  |  | Compartmentalised designed pack that systematically supports pack user in a step by step simple approach | Environmental context and resources | Environmental restructuring  Persuasion  Enablement | 12.5 Adding objects to the environment  The pack design must facilitate pack use rather than reduce it- this should include compartmentalisation of pack components, pack level instructions as well as component level instructions |
|  |  | Links to sites and instructions should be provided which provide further detail of the pack and how to use it | Environmental context and resources  Skills | Environmental restructuring  Persuasion  Enablement | 4.1 Instructions on how to perform a behaviour  On-line materials and links within the pack-level instructions should enable the pack user to access detailed examples of overall pack use and component levels |
| **Pack user reading the information leaflets** | For the pack user, too much text can reduce attention and engagement with the leaflets |  | Memory, attention and decision-making | Environmental restructuring | 12.1 Restructuring the physical environment  The leaflet design must be simple and engaging; it should include some visuals and have no large text blocks  11.3 Conserving mental resources  The leaflet can suggest that the pack user ensures a quiet environment to facilitate reading the leaflet (e.g. turn the TV and radio off before you start to read the leaflets) |
|  | For the pack user an overload of textual information can create problems with attention and focus |  | Memory, attention and decision-making | Environmental restructuring | 12.1 Restructuring the physical environment  The leaflet design must be simple and engaging; it should include some visuals and have no large text blocks  11.3 Conserving mental resources  The leaflet can suggest that the pack user ensures a quiet environment to facilitate reading the leaflet (e.g. turn the TV and radio off before you start to read the leaflets) |
|  | For pack users variations in literacy levels may reduce engagement with the leaflets for some |  | Memory, attention and decision-making | Environmental restructuring | 12.1 Restructuring the physical environment  The leaflet design must be simple and engaging; it should include some visuals and have no large text blocks  11.3 Conserving mental resources  The leaflet can suggest that the pack user ensures a quiet environment to facilitate reading the leaflet (e.g. turn the TV and radio off before you start to read the leaflets) |
|  | Engaging with the leaflets may be difficult for pack users whose first language is not English |  | Memory, attention and decision-making | Environmental restructuring | 12.1 Restructuring the physical environment  The leaflet should be translated into a number of other languages and these can be offered to pack users within the telephone consultation  12.1 Restructuring the physical environment  The leaflet design must be simple and engaging; it should include some visuals and have no large text blocks  11.3 Conserving mental resources  The leaflet can suggest that the pack user ensures a quiet environment to facilitate reading the leaflet (e.g. turn the TV and radio off before you start to read the leaflets) |
|  |  | Articulation of the potential health impact of Chlamydia and other STIs early on within the leaflet may sustain pack user reading and engagement | Beliefs about consequences  Memory, attention and decision-making | Environmental restructuring  Education, persuasion, modelling | 5.1 information about health consequences  The leaflet should provide information very early within its content that highlights the health consequences of effectively treating or not treating chlamydia  5.2 Salience of consequences  Early on within the leaflet the information about the consequences of chlamydia should be designed in such a way as to ensure that it is particularly memorable |
|  |  | Simplified clear visual communication, visual aids and short text extracts would aid pack user reading and engaging with the leaflets | Memory, attention and decision-making | Environmental restructuring | 12.1 Restructuring the physical environment  The leaflet design must be simple and engaging; it should include some visuals and have no large text blocks  11.3 Conserving mental resources  The leaflet can suggest that the pack user ensures a quiet environment to facilitate reading the leaflet (e.g. turn the TV and radio off before you start to read the leaflets) |
|  |  | A sense of the time needed to complete self-sampling should be clearly articulated | Beliefs about consequences  Memory, attention and decision-making | Environmental restructuring  Education, persuasion, | 5.3 information about social and environmental consequences  Provide information within the leaflet about the time needed for completion of the self-sampling. |
| **Correct use of self-sampling kits; swabs, urine and blood samples** | Pack users beliefs about the samples degrading in the post may discourage correct use of self-sampling of self-sampling kits |  | Beliefs about consequences  Knowledge | Education  Enablement, Persuasion, modelling | 5.3 information about social and environmental consequences  On-line pack access, or pack -level instructions must explain the efficacy of the samples for diagnosis when they are distributed through the post |
|  | Pack users beliefs that they will struggle with collecting blood samples may discourage correct use of self-sampling of self-sampling kits |  | Beliefs about Capabilities  Skills | Education, persuasion, modelling, enablement  Training | 15.1 verbal persuasion about capability  On-line pack access, or pack -level instructions can boost the pack user self-efficacy telling them they can do the self-sampling tests; stating that they will succeed  6.1 Demonstration of the behaviour  Within the on-line support there should be a clear observable example of how to perform the self-samples, including the blood samples |
|  | Pack users may be put off the correct use of self-sampling of self-sampling kits by the inclusion of the HIV self-sample kit |  | Knowledge  Social influence  Beliefs about consequences  Emotions | Education  Environmental restructuring  Enablement, Persuasion, modelling | 6.2 Social Comparison  On-line pack access, or pack -level instructions should draw attention to other pack user use of all the pack (e.g., around 90% of pack user successfully return all the self-samples)  5.1 information about health consequences  On-line pack access, or pack -level instructions must explain the rationale for the HIV self-sampling kits and provide a clear articulation of the health consequences of not using it  5.3 information about social and environmental consequences  On-line pack access, or pack -level instructions must explain the rationale for all the HIV self-sampling kits and a clear articulation of the wider consequences of not using it  On-line pack access, or pack -level instructions should provide a sense of choice and partial uptake of the self-samples (HIV or Chlamydia) |
| **Returning the sample** | Pack user perceptions that STI stigma shaped other people’s perceptions of the appearance of the return envelope may reduce willingness to return samples |  | Knowledge  Social influence  Beliefs about consequences  Emotions | Environmental restructuring  Enablement | 12.5 Adding objects to the environment  The return envelope should look like it could be any envelope from a distance and the lab address should be easy to cover with one’s hand  12.5 Adding objects to the environment  Make sure there is a larger envelope available to mask the return envelope with the clinic address |
|  | Pack users perceptions that the appearance of the pack may be read as a diagnosis of Chlamydia may reduce willingness to return samples |  | Knowledge  Beliefs about consequences  Social influence  Emotions | Environmental restructuring  Enablement | 12.5 Adding objects to the environment  The return envelope should look like it could be any envelope from a distance and the lab address should be easy to cover with one’s hand  12.5 Adding objects to the environment  Make sure there is a larger envelope available to mask the return envelope with the clinic address |
|  | Pack users concerns relating to safety and effectiveness of the postal system may reduce willingness to return samples |  | Beliefs about consequences | Education, persuasion, modelling | 5.1 information about health consequences  Pack-level instructions and the site where packs can be accessed on-line should provide clear information concerning the viability of samples which are self-collected, and the viability of samples delivered to labs through the postal system  6.2 Social Comparison  Pack-level instructions and the site where packs can be accessed on-line should draw the pack user s attention to the fact that other people return the samples and get accurate results all the time (‘this is routine care within London clinics’) |
|  | Perceptions that the returned pack might be unsealed or damaged in the post may reduce willingness to return samples |  | Beliefs about consequences | Education, persuasion | 5.1 information about health consequences  Pack-level instructions and the site where packs can be accessed on-line should provide clear information concerning the viability of samples collected and posted  6.2 Social Comparison  Pack-level instructions and the site where packs can be accessed on-line should draw the pack user’s attention to the fact that other people return the samples and get accurate results all the time (‘this is routine care within London clinics’) |
|  |  | Providing a range of return options may increase pack user returning samples back | Memory, attention and decision processes | Environmental restructuring | 12.5 Adding objects to the environment  Pack-level instructions and the site where packs can be accessed on-line should ensure that the pack user knows that there are safe and secure drop off boxes within GUM clinics that represent alternatives to postal delivery |
|  |  | A receipt notification may increase pack user returning samples back | Behavioural regulation | Education, enablement | 2.2 Feedback on behaviour  Systems should be in place that acknowledge receipt of the pack to the pack user.  2.7 Feedback on outcomes of behaviour  Systems should be in place that provide feedback on the outcomes of the use of the self-sampling kits by giving results. Quickly and efficiently including non-reactive results |
